# Psychometric properties of the Copenhagen Burnout Inventory (CBI) in Italian physicians

**DOI:** 10.1101/2022.04.22.22274175

**Authors:** Edoardo Nicolò Aiello, Elena Fiabane, Simona Margheritti, Stefano Magnone, Nadia Bolognini, Massimo Miglioretti, Ines Giorgi

## Abstract

**Background:** Assessing burnout in physicians is relevant as it can adversely affect both their mental and physical health by also decreasing the quality of care, especially since the onset of the COVID-19 pandemic. This study aimed at standardizing the Copenhagen Burnout Inventory (CBI), a psychometrically sound, worldwide-spread tool, in Italian physicians.

**Methods:** Nine-hundred and fifteen Italian physicians were web-administered the CBI, Patient Health Questionnaire-8 (PHQ-8), Generalized Anxiety Disorder-7 (GAD-7) and General Self-Efficacy Scale (GSE). The present CBI is a self-report questionnaire including 18 Likert items (*range*=19-90) assessing Personal, Work-related and Client-related Burnout. Client-related adaptation was performed. Construct validity, factorial structure (Confirmatory Factor Analysis) and internal consistency were tested. Diagnostic accuracy was assessed simultaneously against the PHQ-8, GAD-7 and GSE.

**Results:** All CBI measures yielded optimal internal consistency (Cronbach’s α=.90-.96). The CBI met its original three-factor model (CFI=.94; TLI=.93; RMSEA=.09; SRMR=.04), was positively related with the PHQ-8 (*r*=.76) and GAD-7 (*r*.=73), whereas negatively with the GSE (*r*=-.39), and yielded optimal diagnostics (AUC=.93; sensitivity=.91 and specificity=.85 at the optimal cut-off of 69/90).

**Discussion:** The CBI is a valid, reliable and normed tool to assess burnout levels in physicians, whose use is encouraged in both clinical practice and research as being short-lived, easy to use and openly accessible.

## 1. Background

Burnout syndrome is traditionally defined as a psychological reaction to chronic work-related stress characterized by three dimensions: energy depletion or exhaustion, increased mental distance from one’s job or feelings of negativism or cynicism related to one’s job, and reduced professional efficacy (Schaufeli et al., 2009). This syndrome has an impact on employees’ well-being causing physical weakness, insomnia, anxiety, and depression (Parola et al., 2017), on institutions and systems through expensive job turnover and increased decisional errors, increased absenteeism, and poor work performance (West et al., 2018).

It is thereupon necessary to early detect such a syndrome through ad hoc psychometric tools, in order to aid primary, secondary and tertiary preventive interventions. In this respect, The Maslach Burnout Inventory (MBI; Maslach & Jackson, 1981) has been thoroughly adopted worldwide, but despite acceptable psychometric support, its content validity has been questioned (Kristensen et al., 2005; Schaufeli et al., 2009). In addition, as being copyrighted, the MBI requires organizations to invest fairly large amounts of economic resources.

In such a framework, Kristensen et al. (2005) developed the Copenhagen Burnout Inventory (CBI), a free-to-use tool that extends the construct of burnout syndrome to different domains of workers’ life, as assessing personal, work-, and client-related burnout. Within the CBI, personal burnout is operationalized in terms of feelings of physical, emotional, and mental fatigue and exhaustion, whereas work-related burnout refers to symptoms that respondents attribute to their specific work activity. Client-related burnout instead taps on burnout symptoms selectively referring to respondents’ feelings towards their target clients (e.g., students for teachers, patients for physicians, etc.).

The CBI has been recently standardized in many countries across a wide variety of settings and samples, such as nurses (Montgomery et al., 2021; Thrush et al., 2021), physicians (Lapa et al., 2018; Papaefstathiou et al., 2019; Thrush et al., 2021), pharmacists (Fadare et al., 2021), teachers or university professors (Avanzi et al., 2013; Piperac et al., 2021; Rocha et al., 2020), medical students (Todorovic et al., 2021), and other healthcare employees (Javanshir et al., 2019; Thrush et al., 2021; Walters et al., 2018). In all of these studies, the tool showed good psychometric properties for measuring occupational burnout. Despite this, in Italy, the tool was only validated in teacher samples (Avanzi et al., 2013), although, given its high flexibility in terms of target populations, standardization in other samples has been encouraged by the original Authors themselves (Kristensen et al., 2005).

In this respect, physicians have been historically identified as particularly at-risk for burnout (Rotenstein et al., 2018), and would thus benefit from such an ad hoc standardization study. The latter assertion acquires even greater relevance in the face of the COVID-19 pandemic, which, albeit overall increased the prevalence and incidence of burnout in several occupational settings, undoubtedly poses major pressure especially on healthcare systems, and thus physicians (Baptista et al., 2021; Dehon et al., 2021; Sharifi et al., 2021). The pandemic has indeed entered novel stressors possibly contributing to physician burnout (Amanullah & Shankar, 2020; Bradley & Chahar, 2020): fears of becoming infected or infecting a family member, a lack of appropriate personal protective equipment, impossibility to access to up-to-date information, restricted time with close ones, economic revenue reductions, and increased demands from family responsibilities (Baptista et al., 2021; de Brier et al., 2020; Dehon et al., 2021).

As physician burnout can adversely affect both their mental and physical health and, in turn, decrease the quality of care, it is crucial to assessing their burnout levels through psychometric tools specifically standardized in this populations.

This research thus aims to provide psychometric properties of the CBI among Italian physicians.

## 2. Methods

### 2.1 Participants

Nine-hundred and fifteen licensed physicians (505 females, 410 males; median age class: 41-50 years; median years of service class: 16-25 years) subscribed to the physician’s union ANAAO ASSOMED Associazione Dirigenti Medici, Milano, Italy were e-mailed a web-based questionnaire (Google Form) for data collection, which started on November 21^st^, 2021 and ended on January 14^th^, 2022. Detailed demographic and occupational measures are reported in Supplementary Table 1.

### 2.2 Materials

The Italian CBI by Avanzi *et al*. (2013) is a self-report questionnaire including 19 Likert-like items ranging from 1 (“Never/almost never” for items 1-12 or “To a very low degree” for items 13-19) to 5 (“Always” for items 1-12 or “To a very high degree” for items 13-19); its total score ranges from 19 to 95 (high values corresponding to high burnout levels). Items 1-6 assess Personal Burnout (PB), items 7-10 and 13-15 Work-related Burnout (WB) and items 11-12 and 16-19 Client-related Burnout (CB). For the purposes of this study, the Italian CBI was adapted as follows: (1) items were re-worded by referring to “patients” instead of “students”; (2) item 17 (“Do you find it frustrating to work with clients?”) was dropped as in contrast with physicians’ deontological principles and thus likely to induce social desirability-biased responses. Hence, the present CBI included 18 items and ranges from 5 to 90.

Depression and anxiety were assessed *via* the Patient Health Questionnaire-8 (PHQ-8) (Kroenke et al., 2009) and Generalized Anxiety Disorder-7 (GAD-7) (Spitzer et al., 2006), respectively, whereas self-efficacy *via* the General Self-Efficacy Scale (GSE) (Chen et al., 2001).

### 2.3 Statistics

As skewness and kurtosis values were ≤|1| and ≤|3|, respectively, for all raw psychometric measures, normality and homoscedasticity were assumed as met, and associations of interested were thus tested *via* Pearson’s coefficients (Kim, 2013).

Internal consistency was assessed with Cronbach’s α. Factorial structure was assessed through Confirmatory Factor Analysis (CFA), by addressing the following metrics: root mean square error of approximation (RMSEA), standardized root mean square residual (SRMR), Tucker-Lewis index (TLI), comparative fit index (CFI). Model fit was judged as acceptable according to the following cutoffs (Hu & Bentler, 1999; MacCallum et al., 1996): RMSEA≤.1; SRMR≤.08; TLI and CFI ≥.9.

Diagnostic accuracy of the CBI was tested through receiver-operating characteristics analyses against a state variable defined as the co-occurrence of a PHQ-8 and GAD-7 scores ≥10 (moderate depression and anxiety, respectively) (Kroenke et al., 2009; Spitzer et al., 2006) and a GSE score below the 5^th^ percentile (≤20). The optimal cut-off was identified *via* Youden’s *J* statistics.

In all of the analyses, item 10 was reversed. Multiple comparisons were Bonferroni-corrected where necessary. Analyses were run via SPSS 27 (IBM Corp., 2021) and JASP 0.16.1 (JASP Team, 2022).

## 3. Results

Psychometric measures are summarized in Supplementary Table 2.

The CFA (Figure 1) revealed an optimal fit to the three-factor model (CFI=.94; TLI=.93; RMSEA=.09; SRMR=.04), with all items significantly loading (*p*s<.001) on each CBI subscale (PB: loading *range*=.67-1.02; WB: loading *range*=.59-1.03; CB: loading *range*=.7-1.02). Cronbach’s α was excellent for all CBI measures (Total: .96; PB: .92; WB: .91; CB: .90), with optimal item-total correlations (Total: .51-.84; PB: .74-.88; WB: .59-.83; CB: .56-.83).

**Figure 1.**
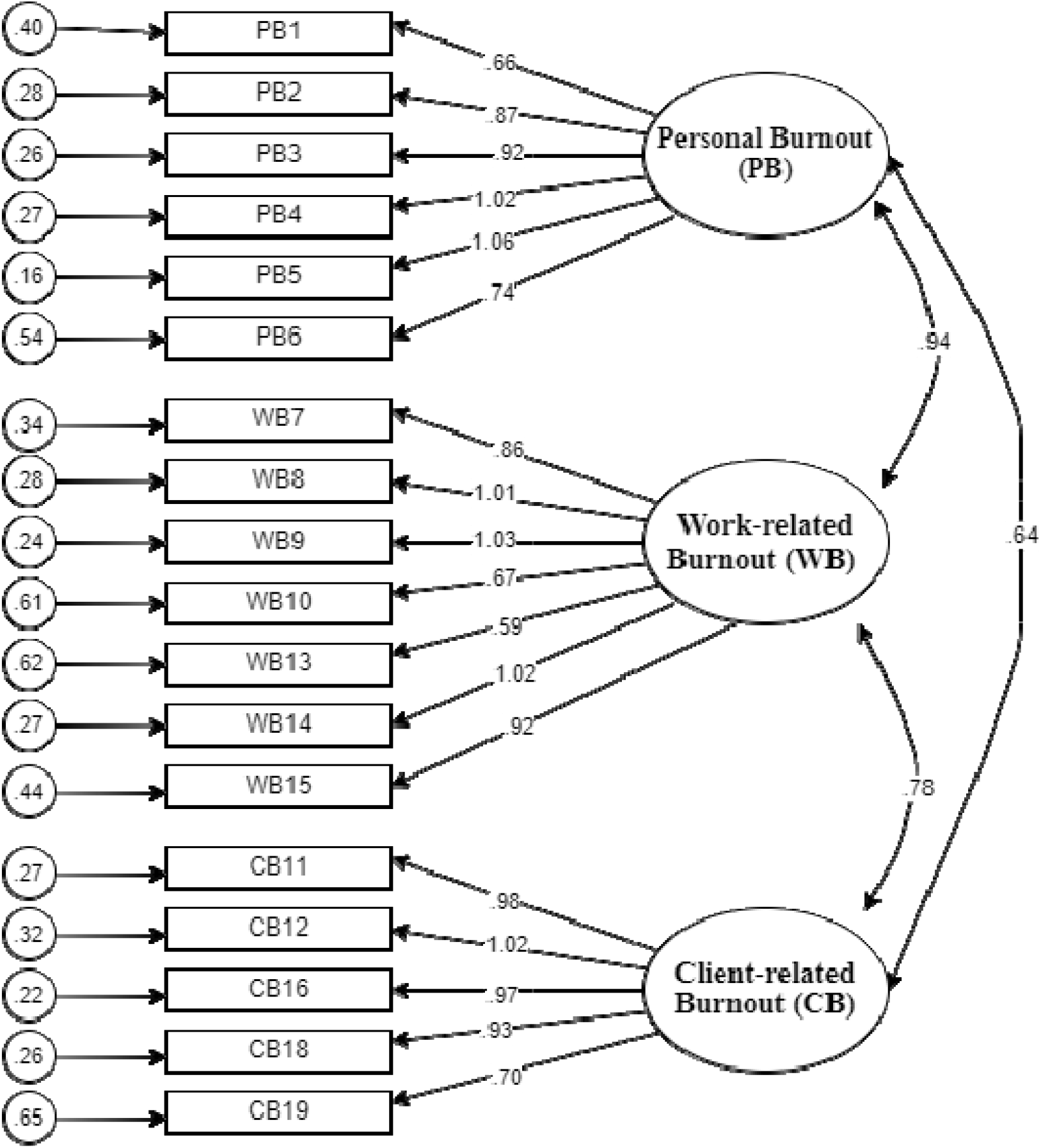
CFA for the CBI in Italian physicians. **Notes**. Rectangles = observed variables; ovals = factors; circles = item uniqueness values; curved arrows = correlation among factors; straight arrows towards observed variables from factors = loadings.

At α_adjusted_ =.017, CBI scores were positively related with the PHQ-8 (*r*(915)=.76; *p*<.001) and GAD-7 (*r*(915)=.73; *p*<.001), whereas negatively with the GSE (*r*(915)=-.39; *p*<.001).

The CBI yielded optimal diagnostic accuracy (AUC=.93; *SE*=.02; CI 95% [.89, .96]) and, at its optimal cut-off (>69/90; *J*=.76), high sensitivity (.91) and specificity (.85). At this cut-off, 16.9% of the sample resulted as presenting with a burnout syndrome.

## 4. Discussion

The present study provides Italian practitioners and researchers with evidence on the psychometric goodness of the CBI as addressed to physicians. The CBI met the three-factor factorial structure originally identified (PB, WB and CB; Kristensen et al., 2005), and all of its measures yielded high internal consistency. Moreover, higher CBI scores were found to be strongly related to higher depression (PHQ-8) and anxiety (GAD-7) levels, as well as moderately with lower self-efficacy (GSE) levels. The latter findings support its both convergent and divergent validity, also filling the existing gaps related to previous CBI standardization which under-addressed such a fundamental psychometric property (Papaefstathiou et al., 2019; Thrush et al., 2021).

It should be also noted that physicians included in the present work came from different types of healthcare organizations (*i.e*., public, private, and academic medical centers), this supporting the external validity of these findings and thus the usability of the CBI in physicians regardless of their specific extraction (Yeh et al., 2018; Yousaf et al., 2021).

This work also provides a cut-off value derived from an empirical algorithm addressing high levels of depression and anxiety and, simultaneously, low levels of self-efficacy, which is both theoretically motivated (Aftab et al., 2012; Denning et al., 2021; Shoji et al., 2015) and supported by the abovementioned data on construct validity. Accordingly, scores on the CBI higher than 69/90 are to be addressed as suggestive of a burnout syndrome and would thus motivate clinical attention. In this respect, the excellent intrinsic diagnostics detected for the CBI at such a cut-off strongly support its clinical use in prevention setting for the early detection of burnout syndrome in physicians.

A limitation should be finally noted, namely that data collection occurred during an ongoing wave of COVID-19 (November 2021-January 2022); hence, greater levels of burnout might have yielded due to such a contingency. However, in this last respect, as being expected that the pandemic will represent a “chronic” burden on healthcare professionals in the next future, the present findings are likely to be likewise representative of the actual *status quo* of burnout syndrome in Italian physicians.

Future research should investigate the measurement invariance of the CBI across representative samples from different Countries. It is indeed encouraged to verify whether it is possible to make meaningful comparisons among physicians’ burnout levels from different Countries, as already done for other burnout tools (Aboagye et al., 2018; de Beer et al., 2020; Reis et al., 2015; Vanheule et al., 2007) The same type of analysis might also be performed in respect to different demographic confounders, as they have been shown to possibly yield heterogeneity in burnout levels (Thrush et al., 2021; West et al., 2018).

In conclusion, the CBI is a valid, reliable, and normed tool to assess burnout levels in physicians for both clinical and research purposes. The good psychometric properties of the CBI herewith reported are consistent with prior research (e.g., Fadare et al., 2021; Javanshir et al., 2019; Kristensen et al., 2005; Lapa et al., 2018; Mahmoudi et al., 2017; Montgomery et al., 2021; Moser et al., 2021; Papaefstathiou et al., 2019; Piperac et al., 2021; Rocha et al., 2020; Thrush et al., 2021; Todorovic et al., 2021; Walters et al., 2018). The CBI is thus suitable for every healthcare organization to obtain information that could help guide both primary and secondary preventive interventions, also considering its short-lived nature and ease of use (by both examiners and examinees’ standpoints).

## Data Availability

All data produced in the present study are available upon reasonable request to the corresponding Author.

## Acknowledgments

the Authors would like to thank Prof. Tage S. Kristensen and Prof. Lorenzo Avanzi for allowing us to adopt the CBI and related materials for the purposes of this study.

---

**Supplementary Table 1.** Participants’ demographic and occupational characteristics.

| Measures (N=915) | Percentage |
| --- | --- |
| **Sex** |  |
| Male | 44.8% |
| Female | 55.2% |
| **Age** |  |
| <30 | 0.9% |
| 31-40 | 24.8% |
| 41-50 | 30.2% |
| 51-60 | 28.4% |
| >60 | 15.7% |
| **Years of service** |  |
| <5 | 6.6% |
| 5-15 | 35.4% |
| 16-25 | 26.6% |
| >25 | 31.5% |
| **Department** |  |
| Clinical immunology | 0.3% |
| Anesthesiology and resuscitation | 11.6% |
| Audiology and phoniatrics | 0.1% |
| Cardiac surgery | 0.8% |
| Cardiology | 7.1 |
| Child neuropsychiatry | 2.4% |
| Clinical pharmacology and toxicology | 0.2% |
| Dermatology and venereology | 0.7% |
| Respiratory system diseases | 2.7% |
| Emergency medicine | 2.4% |
| Endocrinology | 1.5% |
| Dietetics | 0.5% |
| Forensic medicine | 0.2% |
| Gastroenterology | 1.6% |
| General surgery | 7.8% |
| Geriatrics | 1.6% |
| Gynecology and obstetrics | 5.5% |
| Hematology | 1.3% |
| Public health and epidemiology | 0.2% |
| Infectious diseases | 3.4% |
| Internal medicine | 9.5% |
| Maxillofacial surgery | 0.5% |
| Medical genetics | 0.1% |
| Metabolic diseases | 0.2% |
| Microbiology and virology | 0.1% |
| Neonatology | 0.8% |
| Nephrology | 2.8% |
| Neurology | 2.8% |
| Neurosurgery | 1.1% |
| Nuclear medicine | 0.1% |
| Occupational medicine | 1.1% |
| Oncology | 2.6% |
| Ophthalmology | 1.3% |
| Orthopedics and traumatology | 4.3% |
| Otolaryngology | 1.3% |
| Pediatric surgery | 1% |
| Pediatrics | 3.1% |
| Physical medicine and rehabilitation | 1.5% |
| Plastic and reconstructive surgery | 0.2% |
| Psychiatry and clinical psychology | 5.1% |
| Radiodiagnostics | 0.5% |
| Radiotherapy | 0.4% |
| Rheumatology | 0.9% |
| Thoracic surgery | 0.3% |
| Transfusion medicine | 0.8% |
| Urology | 2.2% |
| Vascular surgery | 1.2% |
| Other | 1.7% |
| **Type of contract** |  |
| Fixed-term employee | 1.3% |
| Permanent employee | 94.3% |
| Specializing | 1.4% |

**Supplementary Table 2.**
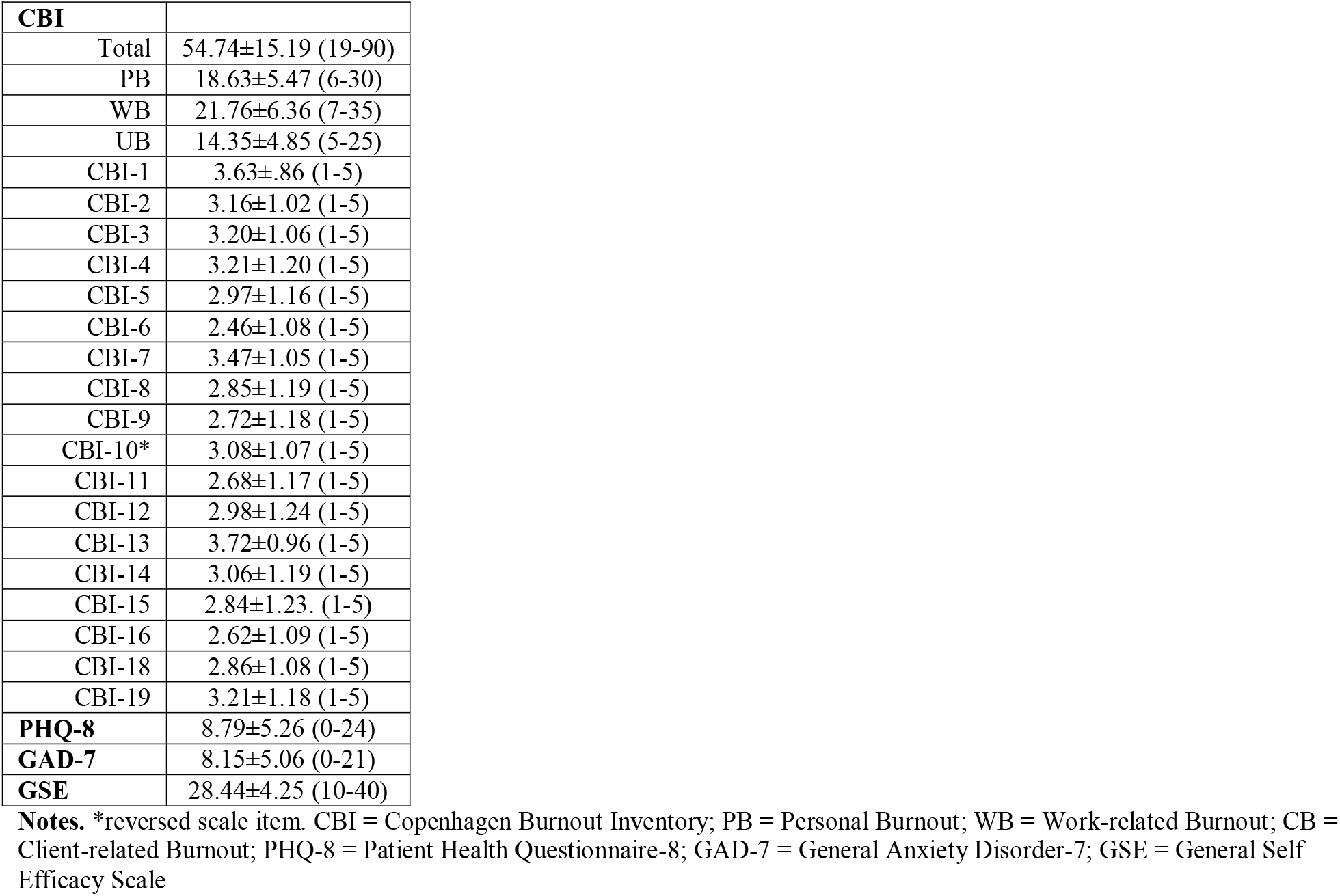
Participants’ psychometric measures.

